# Managing Endothelial Dysfunction in COVID-19: A Pilot, Double-Blind, Placebo-Controlled, Randomized Clinical Trial at the Lebanese American University Medical Center - Rizk Hospital (MEDIC-LAUMCRH)

**DOI:** 10.1101/2022.02.02.22270341

**Authors:** Kamal Matli, Abdulrahman Al Kotob, Wassim Jamaleddine, Soad Al Osta, Pascale Salameh, Rami Tabbikha, Nibal Chamoun, Ahmad Moussawi, Jean-Michel Saad, Gibran Atwi, Tarik Abu Saad, Omar Jamal, Jacques Mokhbat, Georges Ghanem

## Abstract

**Background:** Coronavirus disease 2019 (Covid-19) is associated with endothelial dysfunction. Pharmacologically targeting the different mechanisms of endothelial dysfunction may improve clinical outcomes and lead to reduced morbidity and mortality

**Methods:** In this pilot, double-blind, placebo-controlled, randomized clinical trial we assigned patients who were admitted to the hospital with mild, moderate, or severe COVID-19 infection to receive, on top of optimal medical therapy, either an endothelial protocol consisting of (Nicorandil, L-arginine, Folate, Nebivolol, and Atorvastatin) or placebo for up to 14 days. The primary outcome was time to recovery, measured by an 8 category ordinal scale and defined by the time to being discharged from the hospital or hospitalized for infection-control or other nonmedical reasons. Secondary outcomes included the composite outcome of ICU admission or the need for mechanical ventilation, all-cause mortality, and the occurrence of side effects

**Results:** Of 42 randomized patients, 37 were included in the primary analysis. The mean age of the patients was 57 years; the mean BMI of study participants was 29.14. History of hypertension was present in 27% of the patients, obesity in 45**%**, and Diabetes Mellitus in 21.6%. The median(Interquartile range) time to recovery was not significantly different between the endothelial protocol group (6 [4-12] days) and the placebo group (6 [5-8]days)(p-value = 0.854). Furthermore, there were no statistically significant differences in the need for mechanical ventilation or ICU admission, all-cause mortality, and the occurrence of side effects between the endothelial protocol group and the placebo group.

**Conclusion:** Among patients hospitalized with mild, moderate, or severe COVID-19 infection, targeting endothelial dysfunction by administering Nicorandil, L-arginine, Folate, Nebivolol, and Atorvastatin on top of optimal medical therapy did not decrease time to recovery. However, this treatment protocol was associated with an excellent safety profile. Adequately sized prospective randomized controlled trials are needed for the evaluation of the role of treating endothelial dysfunction in COVID-19 infection.

## Introduction

The coronavirus disease 2019 (COVID-19), caused by the novel severe acute respiratory syndrome coronavirus 2 (SARS-CoV-2), continues to be a global challenge worldwide. Although considered at its emergence as a respiratory infection, recent studies demonstrated tropism towards endothelial cells.^1^ Endothelial dysfunction may be a common pathophysiological process underlying the resulting morbidity and mortality.^2^

Treating endothelial dysfunction is a promising approach to improve outcomes in COVID-19 Patients.^3^ In the resting state, the endothelium regulates blood flow and vascular tone in addition to providing vasodilatory, anticoagulant, anti-inflammatory, and antioxidant properties.^4^ So far, several medications that act directly on the endothelium demonstrated an improved clinical outcome in COVID-19 patients. Dexamethasone and anticoagulants, when employed in the treatment of COVID-19 patients, were found to improve endothelial injury and dysfunction.^5,6^ Clinically, this was translated into a statistically significant decrease in mortality.^7,8^ However, the pathophysiology of endothelial dysfunction is multifactorial and as such can’t be optimally targeted and fully treated with a single agent. This is evidenced by the fact that even with the use of those medications, mortality and morbidity remained high.^4^ Thus, targeting the different mechanisms of endothelial dysfunction concurrently might result in an improved clinical outcome and halt the progression of the disease.

Several medications with endothelial healing properties and excellent safety profiles used for the treatment of other conditions might play a role in the treatment of COVID-19. Nicorandil is a vasodilator drug with antioxidant and anti-inflammatory properties.^9^ Nebivolol, a cardioselective beta-blocker has also shown vasodilating properties, alongside antioxidative,anti-atherosclerotic activities, and antiviral properties.^3^ L-arginine and folic acid, which enhances the synthesis and stabilization of BH4, have also an established role in NO-mediated vasodilation.^10^ Atorvastatin, which is cardioprotective, may also improve endothelial function by increasing the production of NO, and subsequent vasodilatory effect, along with its established anti-inflammatory and antioxidant properties.^3^

Hence, we designed this double-blind placebo-controlled randomized pilot Clinical trial to investigate the efficacy and safety of simultaneously treating the multiple pathways of endothelial dysfunction in hospitalized COVID-19 patients with a 5-drug protocol that consists of Nicorandil, L-arginine, Folate, Nebivolol, and Atorvastatin in an attempt to improve clinical outcome and demonstrate the safety of such a treatment.

## Methods

### Design

This study is a single-center, pilot, double-blind, placebo-controlled, randomized clinical trial (ClinicalTrials.gov Identifier: NCT04631536) at the Lebanese American University Medical Center - Rizk Hospital. The trial protocol was approved by the institutional review board at the Lebanese American University, approval number LAUMCRH.GG2.4/Jan/2021 and was supervised by an independent board responsible for monitoring data safety. Consent was taken from the patient or the patient’s legal guardian in conformity with the IRB rules and regulations.

### Participants

Enrollment for MEDIC-LAUMC began on January 21, 2021, and ended on August 21, 2021, at the Lebanese American University Medical Center - Rizk Hospital (LAUMC-RH). Recruited patients were randomly assigned to receive either the endothelial treatment protocol or a placebo.

### Inclusion Criteria

Patients who were at least 18 years or more, hospitalized with PCR-confirmed COVID-19 with mild, moderate, or severe disease as defined by the U.S Food and Drug Administration, 2020were eligible for inclusion (Table 1).^11^ Moreover, Recruitement had to be done within 48 hours of admission to the hospital.

**Table 1:** Severity Classification of COVID-19 infection

|  |  |
| --- | --- |
| Mild COVID-19 | . Positive COVID-19 testing by standard RT-PCR assay or equivalent test<br>+<br>. Symptoms which could include fever, cough, sore throat, malaise, headache, muscle pain, gastrointestinal symptoms, without shortness of breath or dyspnea<br>. Absence of clinical signs indicative of Moderate, Severe, or Critical disease |
| Moderate COVID-19 | . Positive COVID-19 testing by standard RT-PCR assay or equivalent testing<br>+<br>. Symptoms which could include any symptom of mild illness or shortness of breath with exertion<br>. Clinical signs suggestive of moderate illness with COVID-19, such as respiratory rate $\geq 20$ breaths per minute, saturation of oxygen (SpO<sub>2</sub>) $> 93\%$ on room air at sea level, heart rate $\geq 90$ beats per minute<br>. Absence of clinical signs indicative of Severe or Critical disease |
| Severe COVID-19 | . Positive COVID-19 testing by standard RT-PCR assay or an equivalent test<br>+<br>. Symptoms which could include any symptom of moderate illness or shortness of breath at rest, or respiratory distress<br>. Clinical signs indicative of severe systemic illness with COVID-19, such as respiratory rate $\geq 30$ per minute, heart rate $\geq 125$ per minute, SpO<sub>2</sub> $\leq 93\%$ on room air at sea level or PaO<sub>2</sub>/FiO<sub>2</sub> $< 300$<br>. No criteria for Critical Severity |
| Critical COVID-19 | . Positive COVID-19 testing by standard RT-PCR assay or equivalent test<br>+<br>. Respiratory failure defined based on resource utilization requiring at least one of the following:<br>- Endotracheal intubation and mechanical ventilation, oxygen delivered by high-flow nasal cannula (heated, humidified, oxygen delivered via reinforced nasal cannula at flow rates $> 20$ L/min with fraction of delivered oxygen ≥ |
|  | 0.5), noninvasive positive pressure ventilation, ECMO, or clinical diagnosis of respiratory failure (i.e., clinical need for one of the preceding therapies, but preceding therapies not able to be administered in setting of resource limitation)<br>-Shock (defined by systolic blood pressure < 90 mm Hg, or diastolic blood pressure <60 mm Hg or requiring vasopressors)<br>-Multi-organ dysfunction/failure |

**Table 2:** Patient Characteristics

| Characteristics | All | Placebo | Drug |
| --- | --- | --- | --- |
|  | N=37 | N=20 | N=17 |
| Mean Age - yr | 56.62 | 56.1 | 57.2 |
| Male Sex - No. (%) | 30 (81.8) | 17 (85) | 13 (76.5) |
| Female Sex - No. (%) | 7 (18.9) | 3 (15) | 4 (23.5) |
| BMI | 29.14 | 29.12 | 29.9 |
| Days since symptom onset | 7.6 | 6.78 | 8.56 |
| SpO2 on arrival - % | 91.6 | 91.2 | 92.1 |
| Coexisting Conditions - No./Total No. (%) |  |  |  |
| Hypertension | 10 (27) | 7 (35) | 3 (17.6) |
| Diabetes Mellitus | 8 (21.6) | 6 (30) | 2 (11.8) |
| Coronary Artery Disease (CAD) | 2 (5.4) | 2 (10) | 0 (0) |
| Congestive Heart Failure (CHF) | 0 (0) | 0 (0) | 0 (0) |
| Asthma | 4 (10.8) | 2 (10) | 2 (11.8) |
| Chronic Obstructive Pulmonary Disease (COPD) | 0 (0) | 0 (0) | 0 (0) |
| Cancer | 1 (2.7) | 0 (0) | 1 (5.9) |
| Chronic Kidney Disease (CKD) | 1 (2.7) | 1 (5) | 0 (0) |
| Thrombo-embolic Disease | 0 (0) | 0 (0) | 0 (0) |
| Current Tobacco Use | 10 (27) | 5 (25) | 5 (29.4) |
| Previous Tobacco Use | 2 (5.4) | 0 (0) | 2 (11.8) |
| Alcohol Use | 4 (10.8) | 2 (10) | 2 (11.8) |
| Previous Medications Used - No. (%) |  |  |  |
| ACEI or ARB | 6 (16.2) | 4 (20) | 2 (11.8) |
| Anticoagulation | 14 (37.8) | 8 (40) | 6 (35.3) |
| Antiplatelets | 14 (37.8) | 10 (50) | 4 (23.5) |
| Corticosteroids | 18 (48.6) | 12 (60) | 6 (35.3) |
| Immunosuppressants | 1 (2.7) | 0 (0) | 1 (5.9) |
| Colchicine | 3 (8.1) | 2 (10) | 1 (5.9) |
| Ivermectin | 8 (21.6) | 2 (10) | 6 (35.3) |
| Inpatient Treatments during trial - No. (%) |  |  |  |
| Supplemental Oxygen | 31 (83.8) | 17 (85) | 14 (82.4) |
| Glucocorticoids | 34 (91.9) | 19 (95) | 15 (88.2) |
| Remdesivir | 22 (59.5) | 12 (60) | 10 (58.8) |
| Favipiravir | 5 (13.5) | 3 (15) | 2 (11.8) |
| Tocilizumab | 3 (8.1) | 3 (15) | 0 (0) |
| Tofacitinib | 15 (40.5) | 7 (35) | 8 (47.1) |
| Colchicine | 5 (13.5) | 2 (10) | 3 (17.6) |
| Anticoagulation | 37 (100) | 20 (100) | 17 (100) |
| Antiplatelets | 15 (40.5) | 12 (60) | 3 (17.6) |

### Exclusion criteria

Exclusion criteria for recruitment include patients with any of the following: Patients with critical covid-19(Table 1). In addition, other exclusion criteria included inability to tolerate oral medications, patients already taking beta-blockers, Nicorandil, PDE5 inhibitors, or Riociguat, patients with shock defined by SBP<90 mmHg for more than 30 minutes not responding to IV fluids with evidence of end-organ damage, severe bradycardia (<50 bpm) or heart block greater than first-degree (except in patients with a functioning artificial pacemaker), decompensated heart failure and sick sinus syndrome (unless a permanent pacemaker is in place), severe hepatic impairment (Child-Pugh class C) or active liver disease with unexplained persistent elevations of serum transaminases, pregnant or breastfeeding patients, hypersensitivity to any of the medications in the trial, patients who took the protocol for less than 48 hours, myocarditis, acute pulmonary edema, hypovolemia, patients enrolled in a different randomized study and/or COVID-19 interventional study.

### Randomization Procedures and study interventions

The hospitalized patients enrolled in the trial received the medications or placebo daily up to 14 days or until hospital discharge or death, whichever event occurred first. At the hospital, the study participants were assessed daily from day 1 until day 28. Patients discharged before 28 days were assessed by telephone once weekly. All adverse events or hypersensitivity reactions were recorded.

Assignment of patients was done in a 1:1 ratio between the endothelial protocol arm or the placebo arm. A staff member who had no role in the study managed randomization and created the randomization list via a computer-generated code with block sizes of 8 and sent directly to the central hospital pharmacy where the kits were prepared by pharmacy personnel not included in the trial.

In the endothelial protocol group, patients received a 5-drug regimen. Atorvastatin 40 mg per os once daily was given to statin naïve patients. However, patients already taking a statin as part of their home treatment, had the same statin resumed because it was unethical to remove such therapy with the possibility of having a placebo given instead after preliminary reports showed a clinical benefit might be associated with the use of statins in COVID-19 patients.^12^ In addition, Folic acid and L-arginine were given per os as 5 mg once daily and 1 g 3 times daily, respectively. Nicorandil was given at a dose of 10 mg twice daily. Nebivolol was given at doses of 2.5mg or 5mg PO once daily. The dose was chosen based on the patients’ heart rate upon trial enrollment where patients with higher heart rates received the 5 mg dose. In the placebo group, patients received a matching 5-drug placebo regimen that contained an identical number of pills administered similarly at the same schedule as the endothelial treatment arm. In the case where the patient continued the statin taken at home, 1 less placebo pill was administered. The medications were prepared and labeled by the central pharmacy staff who did not participate in the study. Patients and investigators were blinded to randomization throughout the study period up until the final analysis.

**Figure 1:**
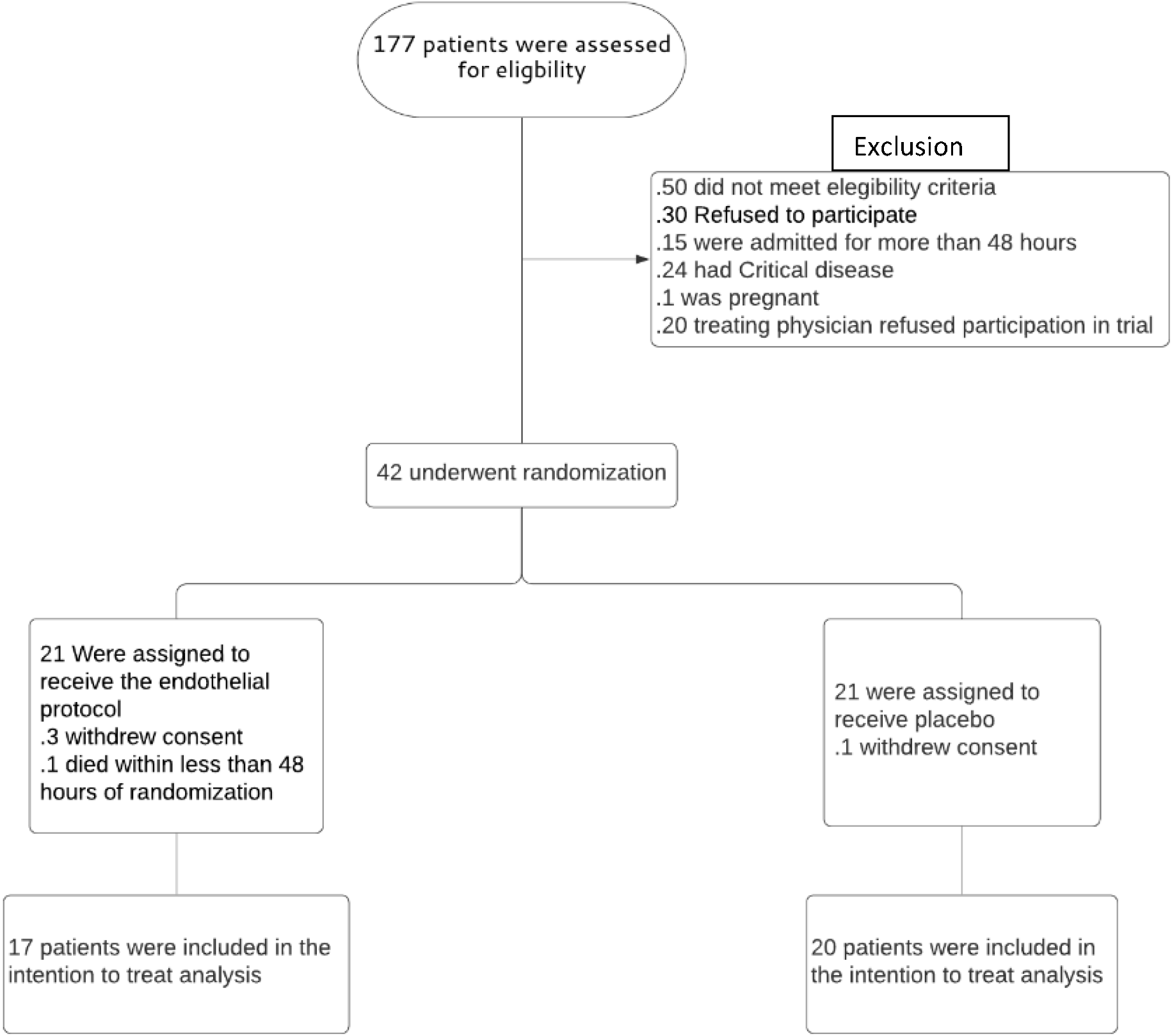
Consort diagram of the study.

### Outcomes

The primary outcome was the time to recovery, defined as the first day, during the 28 days after enrollment, on which a patient met the criteria for category 1, 2, or 3 on the eight-category ordinal scale. The categories are as follows: 1, not hospitalized and no limitations of activities; 2, not hospitalized, with limitation of activities, home oxygen requirement, or both; 3, hospitalized, not requiring supplemental oxygen and no longer requiring ongoing medical care (used if hospitalization was extended for infection-control or other nonmedical reasons); 4, hospitalized, not requiring supplemental oxygen but requiring ongoing medical care (related to COVID-19 or other medical conditions); 5, hospitalized, requiring any supplemental oxygen; 6, hospitalized, requiring noninvasive ventilation or use of high-flow oxygen devices; 7, hospitalized, receiving invasive mechanical ventilation or extracorporeal membrane oxygenation (ECMO); and 8, death.

The secondary outcomes were: the need for invasive mechanical ventilation or ICU admission, all-cause mortality, and occurrence of side effects.

### Statistical Analysis

Calculations were done using the G*Power software.. The time to clinical improvement, which is the primary outcome, was estimated to be approximately 10 days, alpha was fixed at 5% and beta at 20%; subsequently, the sample size was calculated. At least 17 patients in each group and minimum total sample size for both arms=34 were expected to make a difference of 4 days in time to clinical improvement, with a power of 80%. So the study was stopped early when those numbers were achieved.

Data was entered on Excel and analyzed using SPSS version 25.0. A descriptive analysis presented the data, using means and standard deviation for normal continuous variables, median and interquartile range for non-normal variables, while frequency and percentages were used for multinomial and dichotomous variables. For the bivariate analysis, we used the chi-square test to compare percentages, and the COX regression to compare time to requiring mechanical ventilation or ICU admission. In all cases, a p-value <0.05 was considered significant.

## Results

### Patients

177 patients were assessed for eligibility. 42 were eligible and randomized to either the endothelial protocol or the placebo group. The rest were not eligible for inclusion due to the following reasons: 50 patients did not fulfill the inclusion or exclusion criteria, 30 patients refused to participate, and 15 patients were missed by recruiters since they were in the hospital for more than 48 hours when assessed for eligibility. Moreover, 24 already had critical disease, 1 was pregnant and 20 had their treating physicians refusing participation in the trial. Of the 42 randomized patients, 4 withdrew their consent and 1 died after less than 48 hours of randomization and thus were excluded.

A total of 37 were included in the final analysis and underwent randomization (17 to the endothelial protocol group and 20 to the placebo group). They were included in the primary intention-to-treat analysis, received the corresponding study protocol, and were followed up for a total of 28 days.

The baseline characteristics of the patients are presented in table 1. No significant differences in vital signs, laboratory results, or baseline medications were observed between groups at baseline (p>0.05 for all). The mean age of the study participants was 57 years. There was a higher percentage of male patients in this study (81.1%). The mean BMI of study participants was 29.14. History of hypertension was present in 27% of the patients, obesity in 45**%**, and Diabetes Mellitus in 21.6%. The average number of days since symptom onset was 7.6 days.

### Primary Outcome

The median (Interquartile range) time to recovery was not significantly different between the endothelial protocol group (6 [4-12] days) and the placebo group (6 [5-8] days) (p-value = 0.854). (Table 3)

**Table 3:** Primary and Secondary outcomes

|  | Endothelial Protocol | Placebo | P-value |
| --- | --- | --- | --- |
| **Primary Outcome** |  |  |  |
| Time to Recovery, days-<br>median[IQR] | 6 [4-12] | 6 [5-8] | 0.854 |
| **Secondary Outcomes** |  |  |  |
| Need for mechanical<br>ventilation or ICU<br>admission-n (%) | 3 (17.6%) | 2 (10%) | 0.644 |
| All-cause Mortality-n<br>(%) | 0 (0%) | 0 (0%) | – |
| Adverse Events – n (%) | 3 (17.6%) | 3 (15%) | 0.828 |

### Secondary outcomes

There were no statistically significant differences in the need for mechanical ventilation or ICU admission between the endothelial protocol group and the placebo group (17.6% vs. 10%; OR=1.93[95% CI,0.28-13.16]; P= 0.644.. Furthermore, during the 28-day follow-up, no mortality cases were recorded in either arm. However, after the 28 days follow-up duration, 1 patient from each arm eventually passed away.(Table 3)

### Safety Outcomes

All patients were followed up for a period of 28 days. The use of the endothelial protocol was not associated with an increased risk of the overall incidence of side effects when compared to placebo (Odd’s Ratio=1.213; 95% CI 0.211 to 6.985, p=0.828). There was a total of 6 adverse events (3 in the endothelial protocol and 3 in the placebo group) experienced by 6 different patients. There were no severe adverse events. (Table 3)

The most common side effect was bradycardia, defined as having a heart rate less than 50 bpm, which was responsible for 4 out of the 6 reported side effects. (3 in the endothelial protocol mainly when the dose of 5 mg of nebivolol was used and 1 in the placebo arm). The remaining events were 1 case of nausea and vomiting which was in the placebo group and one case of hypotension in the endothelial protocol group where Nebivolol 5 mg was stopped and restarted at a dose of 2.5 mg the next day with no further problems recorded.

## Discussion

To the best of our knowledge, this is the first pilot, double-blind, placebo-controlled, randomized clinical trial to assess the efficacy and safety of managing endothelial dysfunction in COVID-19 patients. In our study, targeting endothelial dysfunction medically did not result in a statistically significant clinical benefit or reach a statistically significant difference in the primary endpoints and the secondary outcomes among hospitalized patients with non-critical COVID-19 infection. However, the endothelial protocol demonstrated an excellent safety profile.

Covid-19, in the end, in its acute form or as long covid syndrome is a disease of the endothelium.^4,13,14^ Thus, and as recommended by the European Society of Cardiology (ESC), we therapeutically targeted the different mechanisms of endothelial dysfunction. So, in addition to optimal medical therapy against COVID-19 infection, we used a 5-drug endothelial protocol with proven efficacy against endothelial dysfunction and an excellent safety profile. This protocol consisted of Nicorandil, L-arginine, Folic Acid, Nebivolol, and Atorvastatin.

Several studies showed a positive outcome when endothelial dysfunction was targeted in COVID-19 patients. Khider et. Al conducted a prospective cohort and reported that COVID-19 patients treated with anticoagulation prior to admission demonstrated a statistically significant decrease in the circulating endothelial cells (CECs), a marker of endothelial injury and dysfunction when compared to those who were untreated(p=0.02).^15^ Furthermore, Targeting the endothelial activation by monoclonal antibodies Narsoplimab and Adrecizumab were associated with improved outcomes in COVID-19 patients.^16,17^ The clinical benefits noted with the use of Tocilizumab and Dexamethasone were attributed to the immunomodulation at the level of the endothelium.^18,19^ Byttebier et. al demonstrated in a retrospective study that the concurrent treatment with statins, aspirin, and ACE/ARBs, which have protective effects on the endothelium, resulted in a 3-fold reduction in the odds of hospital mortality.^20^ In our study, treatment with the endothelial dysfunction protocol resulted in no benefit probably due to the small sample size and the fact that the meantime after presentation was around 7.5 days. By then, the acute phase had already passed and the endothelial damage incurred probably became irreversible with permanent target organ damage. Furthermore, the vast majority of patients received a treatment with anticoagulation and steroids. As mentioned previously, those medications have a proven efficacy in treating endothelial dysfunction in COVID-19 and as such might have blunted the therapeutic response to our protocol keeping in mind the small sample size. In addition, approximately half of the patients received antibiotics which can alter the microbiome by that impacting the inflammatory response.

Even though treating endothelial dysfunction did not demonstrate a better clinical outcome, it showed an excellent safety profile with no increase in the occurrence of side effects. Thus, exploring the effects and outcomes of treating of endothelial dysfunction should be done in adequately sized clinical trials due to the major importance endothelial dysfunction holds as a therapeutic target for the treatment of COVID-19 infection.

The strengths of this study include the randomized, double-blind, placebo-controlled, experimental design; it was done in a day-to-day health-care setting. Furthermore, an extensive safety analysis as well as extensive adverse event reporting were done.

This pilot has several limitations that warrant careful interpretation of the results. The relatively low sample size in this trial could have had inadequate power to exclude small, but clinically meaningful findings, since it was designed to explore a potential signal for the use of endothelial protocol in COVID-19, not to provide definitive evidence on the subject. This pilot was restricted to subjects with non-severe disease. The patients had several coexisting diseases and were subjected to a diverse medication regimen. Consequently, larger scale studies are thus necessary to demonstrate efficacy and confirm safety of the suggested protocol.

## Conclusion

Among hospitalized patients with COVID-19, the use of an endothelial protocol consisting of Atorvastatin, L-arginine, Folic Acid, Nebivolol, and Nicorandil to treat endothelial dysfunction did not significantly improve clinical outcomes. However, an excellent safety profile was found. Thus, adequately sized prospective randomized controlled trials are needed for the evaluation of the role of treating endothelial dysfunction in COVID-19 infection.

## Data Availability

All data relevant to the study are included in the article and are available on reasonable request from the last author GG

## Contributors

KM contributed to hypothesis conception, study design, proposal preparation, patient recruitment and follow-up, data analysis, and interpretation, and wrote the final draft of the manuscript. AK, WJ, SO and GA contributed to patient recruitment and follow-up and contributed to writing the manuscript. PS contributed to data analysis and interpretation as well as critical revisions of the manuscript. JMS and AM contributed to proposal preparation patient recruitment and follow-up. TAS and OJ contributed to patient recruitment and follow-up. NC JM and GG supervised the progress of the trial and provided scientific feedback as well as critically reviewed the manuscript. All authors read and approved the final version of the manuscript before submission for publication.

## Acknowledgments

We thank Dr. Marwan Malek and PharmM pharmaceutical manufacturing company for providing us with the placebo pills and L-arginine. We thank Mrs. Amira Al Kotob for designing the Trial Logo. We thank Mr. Christian Sawma and the Central Pharmacy personnel for medication kit preparation. We Thank Mr. Sami Rizk for approving the use of the trial medications. We thank Dr. Christy Costanian for her input concerning statistics and initial study organization.

## Funding

Trial Medications were provided by the Lebanese American University Medical Center Rizk Hospital

## Competing interests

None

## Notes

### Competing Interest Statement

The authors have declared no competing interest.

### Clinical Trial

NCT04631536

### Author Declarations

institutional review board at the Lebanese American University

